# Pramipexole Enhances Reward Learning by Preserving Value Estimates

**DOI:** 10.1101/2022.01.14.22269287

**Authors:** Don Chamith Halahakoon, Alexander Kaltenboeck, Marieke Martens, John G. Geddes, Catherine J. Harmer, Philip Cowen, Michael Browning

## Abstract

**Background:** Dopamine D2-like receptor agonists show promise as treatments for depression. They are thought to act by altering how individuals learn from rewarding experiences. However, the nature of these reward learning alterations, and the mechanisms by which they are produced is not clear. Reinforcement learning accounts describe three distinct processes that may produce similar changes in reward learning behaviour; increased reward sensitivity, increased inverse decision temperature and decreased value decay. As these processes produce equivalent effects on behaviour, arbitrating between them requires measurement of how expectations and prediction errors are altered. In the present study, we characterised the behavioural effects of a sustained 2-week course of the D2/3/4 receptor agonist pramipexole on reward learning and used fMRI measures of expectation and prediction error to assess which of these three mechanistic processes were responsible for the behavioural effects.

**Methods:** 40 healthy volunteers (Age: 18-43, 50% female) were randomly allocated to receive either two weeks of pramipexole (titrated to 1mg/day) or placebo in a double-blind, between subject design. Participants completed a probabilistic instrumental learning task, in which stimuli were associated with either rewards or losses, before the pharmacological intervention and twice between days 12-15 of the intervention (once with and once without fMRI). Both asymptotic choice accuracy, and a reinforcement learning model, were used to assess reward learning.

**Results:** Behaviourally, pramipexole specifically increased choice accuracy in the reward condition, with no effect in the loss condition. Pramipexole increased the BOLD response in the orbital frontal cortex during the expectation of win trials but decreased the BOLD response to reward prediction errors in the ventromedial prefrontal cortex. This pattern of results indicates that pramipexole enhances choice accuracy by reducing the decay of estimated values during reward learning.

**Conclusions:** The D2-like receptor agonist pramipexole enhances reward learning by preserving learned values. This is a plausible candidate mechanism for pramipexole’s observed anti-depressant effect.

## Introduction

Major depressive disorder (MDD) is a debilitating condition and a pressing public health concern(1). The majority of depressed individuals respond only partially to treatment and a sizable proportion do not respond at all (2), motivating the search for novel interventions. The success of this search depends on characterising promising treatment targets associated with the illness, and identifying agents able to engage these targets. A target of particular recent interest is the impairment of reward learning seen in depressed patients (3,4). The consistent finding in this patient group is that they respond less reliably than control participants to stimuli which are associated with rewards (5–16).

Computational characterisation using reinforcement learning (RL) models has identified three distinct alterations of learning and decision-making process that may produce the behaviour observed in depressed patients (Figure 1b-c): they may make decisions less deterministically (9), they may treat rewards “as if” they were of reduced value (9), or their learned value estimates may decay to a greater degree over time (17).

**Figure 1:**
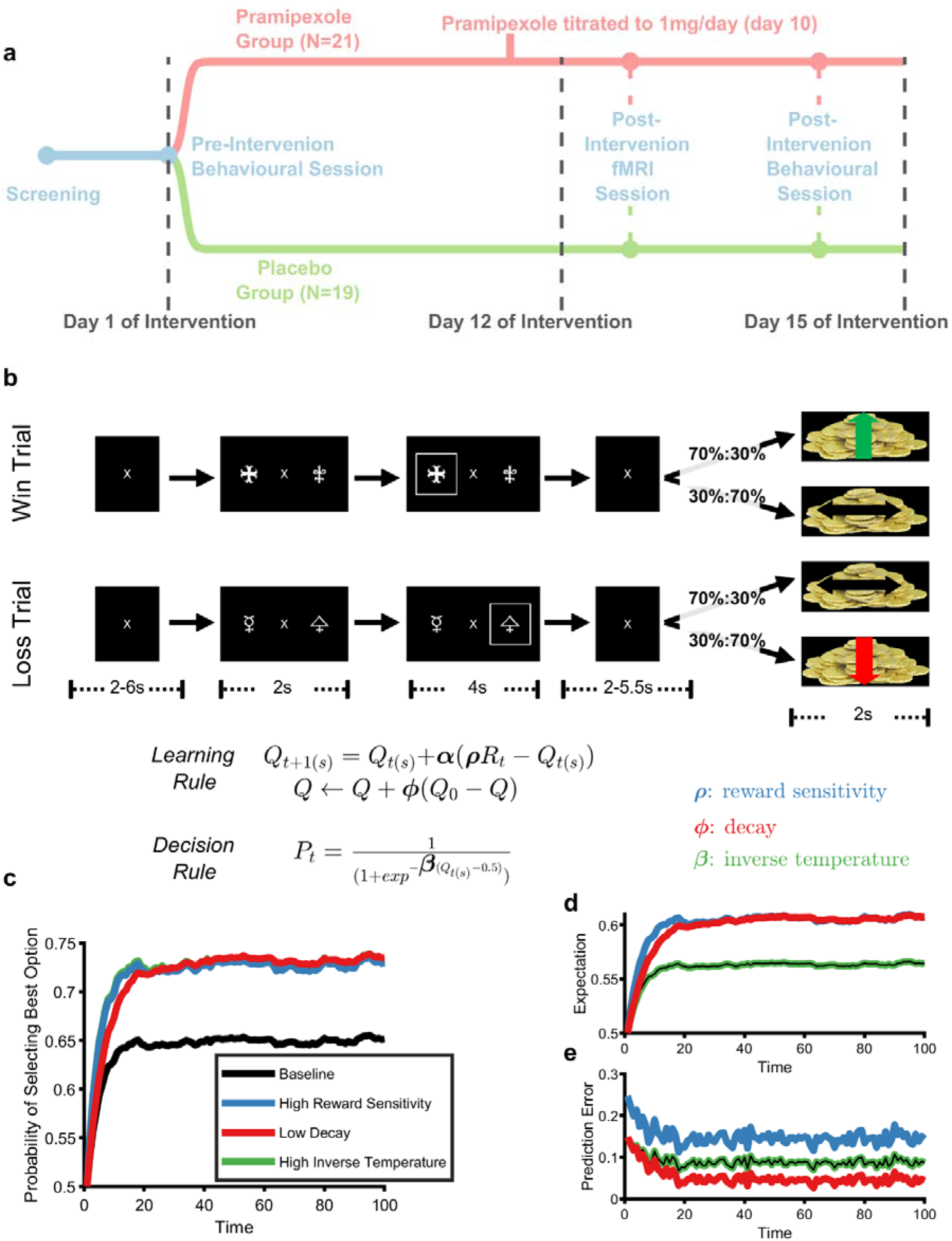
Panel **a**, study design. Following a screening session, participants underwent the pre-intervention behavioural testing session in which they performed the PILT task (described in panel **b**). Participants received the first dose of pramipexole/placebo at the end of this behavioural testing session. Between days 12-15 of the pramipexole/placebo course, participants attended an fMRI session (in which they performed the PILT whilst undergoing fMRI) and, on a separate day, a behavioural testing session which was identical to the pre-intervention behavioural testing session. Panel **b**. Probabilistic Instrumental Learning Task (PILT). In each trial, participants were presented with one of two possible pairs of shapes. For one of the shape pairs (top line), one shape was associated with winning money on 70% of trials and not winning on the remaining 30% (the other shape had reciprocal contingencies). For the other shape pair (bottom line), one shape was associated with losing money on 70% of trials and not losing on the remaining 30% (again, the other shape had reciprocal contingencies). Participants had to learn to select the shapes that were associated with the high probability of win/no-loss. Depression is associated with reduced asymptotic choice of rewarding outcomes in this and similar tasks and so we hypothesized that pramipexole would have the opposite effect (i.e. increase asymptotic choice). Performance on the task can be described by a simple learning rule, in which rewards, *R*, are combined with expectations, *Q*, before being fed into a decision rule. Distinct parameters modify each component of this process: a reward sensitivity parameter, *ρ*, influences the effective size of experienced rewards, a decay parameter, *ϕ*, influences the degree to which expectations are maintained between trials, a learning rate parameter, *α*, influences the rate at which rewards alter expectations and an inverse temperature parameter, *β*, influences the degree to which expectations are used to determine choices. Panel *c*. Learning curves generated by the learning and decision-making rules described in panel b. Choices of the baseline model (black line) were produced using a *ρ* of 0.6, a *ϕ* 0.12, a *β* of 10 and an *α* of 0.1. Increases in either *ρ* (blue line) or *β* (green line) and decreases in *ϕ* (red line) produce equivalent changes in asymptotic choice. In other words, three qualitatively distinct processes lead to the same behavioural effect. As a result, choice data on its own cannot be used to distinguish between these processes. However, the internal model variables do differ, and thus can discriminate, between these processes. Panel **d** illustrates model expectations, *Q*. As can be seen, either increasing *ρ* or decreasing *ϕ* causes an increase in expectations, whereas *β*, which influences decision-making rather than learning, does not change expectations (i.e. the green and black lines are identical). Panel **e** illustrates the prediction errors (PE) of the models, which are able to fully discriminate the three parameters. Again, changes in *β* have no effect, whereas increases in *ρ* leads to increased Pes and reductions in *ϕ* leads to decreased PEs. In order to discriminate between the three possible causes of changed asymptotic choice behaviour, estimates of the internal model variables, such as those produced by neuroimaging measures, are required.

While these processes describe qualitatively distinct causal mechanisms, they produce very similar effects on behaviour in the commonly used reward-learning tasks and thus cannot be distinguished using behavioural outcomes from these tasks alone (9). Rather, measures of the internal model variables that do differ between the processes, such as the neural response to expectations or reward prediction errors, are required (see Figure 1d-e). Neuroimaging measures of striatal and reward-related cortical regions provide an index of these processes (18,19), and thus can help to distinguish between competing mechanistic hypotheses (18,20). The most common neuroimaging finding in depressed and anhedonic populations is a reduced neural response to rewarding outcomes and/or reward prediction errors (7,21–24), suggesting that patients respond less consistently to rewards because they treat outcomes as if they were less rewarding (7), rather than due to decreased decision consistency (which should not affect the BOLD response to reward) or increased value decay (which should increase BOLD response to reward, reflecting greater disparity between reward-expectation and reward-outcome resulting from increased decay of the former).

The centrality of the mesocorticolimbic dopaminergic system in reward learning (25) suggests that dopaminergic agents might act to reverse impaired reward learning in patients with MDD (26). Early evidence suggests that one such agent, the D2-like (D2, D3 and D4) receptor agonist pramipexole, is efficacious in the treatment of MDD (27–29). However, contrary to its clinical effects, previous experimental studies (30–36) of pramipexole generally indicate that it blunts rather than enhances participants’ behavioural responses to reward (30–33). Similarly, pramipexole has been found to blunt the neural response to positive outcomes in reward sensitive brain regions such as the medial prefrontal cortex (mPFC), orbitofrontal cortex (OFC)(37), ventral striatum (VS) and midbrain (38). One explanation for the seeming contradiction between the clinical and experimental evidence is that experimental studies have predominantly examined the effect of a single dose of pramipexole while sustained treatment is required to improve symptoms (27,39). From a pharmacological perspective, acute treatment with D2/3/4 agonists are believed to primarily influence inhibitory presynaptic auto-receptors, leading to reduced dopaminergic transmission, whereas sustained administration leads to auto-receptor down-regulation and enhanced transmission via agonism at post-synaptic D2-like receptors (40–43). This suggests that the clinically relevant effects of pramipexole on reward learning are likely to become apparent only after sustained administration of the drug.

In the present study, we examined the effect of a sustained (2-week) course of pramipexole on both behavioural and neural measures of reward learning (Figure 1a) in non-clinical participants. As registered in clinicaltrials.gov (https://clinicaltrials.gov/ct2/show/NCT03681509), we hypothesized that pramipexole would induce the opposite pattern of reward-learning behaviour characteristic of depression and anhedonia, increasing asymptotic choice of stimuli associated with higher levels of reward (Figure 1c) by increasing subjective valuation of rewards (leading to increased BOLD response to rewarding outcomes in the brain’s reward prediction error network) (44).

## Methods

### Participants, Design and Intervention

We conducted a randomized, placebo controlled experimental medicine study with a between-groups design. 42 non-clinical participants, between the age of 18 and 45, were randomized 1:1 to receive pramipexole or placebo. Potential participants were excluded if they had ever been diagnosed with a psychiatric illness (determined using the SCID-V-CV) or had a first degree relative with a psychotic illness, were taking psychoactive medication, had any history of impulse control difficulties, had any contraindication to pramipexole, had taken any recreational drugs in the last three months, regularly drank more than 4 units of alcohol per day, smoked more than 5 cigarettes per day or drank more than 6 caffeinated drinks per day. Female participants who were pregnant, lactating, or not using a highly effective method of contraception were also excluded. From a starting dose of 0.25mg of pramipexole salt, the dose was increased in 0.25mg increments every 3 days, reaching a dose of 1mg/day by day 10. Participants continued to take 1mg/day for 3-5 days (until testing was completed). Following this, the dose was down-titrated over 3 days to avoid withdrawal effects. The apparent dose of the placebo was increased in the same manner. Participants performed a probabilistic instrumental learning task (PILT; see below for details) before the intervention and then twice between days 12-15 of the intervention (one with fMRI data collection, one behavioural). At the screening session, participants completed the Eysenck Personality Questionnaire (EPQ), Becks Depression Inventory (BDI) and Spot-the-word test (an estimate of IQ). At both behavioural testing sessions, participants completed the Befindlichkeitsskala (BFS), Positive and Negative Affect Schedule (PANAS), State-Trait Anxiety Inventory (STAI), Snaith-Hamilton Pleasure Scale (SHAPS), Temporal Experience of Pleasure Scale (TEPS), Oxford Happiness Questionnaire (OXH) and Impulsive-Compulsive Disorders in Parkinson’s Disease-Rating Scale (QUIP). At the post-intervention behavioural testing session, participants additionally completed a side-effects questionnaire. Two participants (both in the placebo group) dropped out of the study due to nausea.

### Task

Participants completed a modified version of a probabilistic instrumental learning task (PILT) described by Pessiglione et al(18). The PILT is a 2-arm bandit task (Figure 1a) with interleaved ‘win’ and ‘loss’ trials. In each trial, the participant is presented with two stimuli which have reciprocal probabilities (0.7 *vs* 0.3) of a ‘win’ outcome (+£0.20) *vs* a ‘no win’ outcome (£0.00) in reward condition trials, or a ‘loss’ outcome (-£0.20) *vs* a ‘no loss’ outcome (£0.00) in loss condition trials. Participants choose one of the two stimuli, following which they received visual feedback on the trial outcome and their current total earnings. Each block of the PILT consists of 30 reward trials and 30 loss trials. Participants performed 3 blocks of the PILT in each behavioural testing session and 2 blocks in the imaging session. Different task stimuli were used in each block. Participants started each session with £1.50 of funds. Participants received a portion of their winnings from these tasks (up to a maximum of £30).

The behavioural measure of interest was choice accuracy, defined as the proportion of advantageous choices made i.e. the stimulus with 0.7 probability of ‘win’ in the reward condition or the stimulus with 0.7 probability of ‘no-loss’ in the loss condition. We measured accuracy in the second half of each block as this provides a close estimate of asymptotic choice (45,46) found to be associated with depression (see Figure 1c). Note, the same pattern of results were found in the current study if accuracy was calculated across all trials rather from than those in the second half (see supplementary material).

### Reinforcement Learning Models

We used a simple reinforcement learning model, which combined parameters from different, previously described models, to formalize the mechanistic question being addressed in this study.

First a learning rule was used to update expectations about the association stimuli with the outcomes:

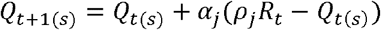

Here, *Q*_*t*(*s*)_ is the expectation about the value of shape *s* on trial *t, R*_*t*_ is the observed outcome (1 for positive outcome, 0 for negative outcome), *α*_*j*_ is the learning rate used for trial valence *j* (i.e. win or loss trial) and *ρ*_*j*_ is a reward sensitivity parameter for trial valence *j*. Expectations were initialized at *Q*_O(*s*)_ *= 0*.*5*, and the unchosen option, *Q*_*t*(*s*′)_, was updated with the reciprocal outcome (see supplementary materials for evidence supporting these decisions). Following this, the model’s expectations decayed back towards the initial value with the rate of decay controlled by a decay factor *ϕ* (17):

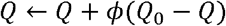

Finally, the *Q* values were fed into a softmax action selector to produce a choice:

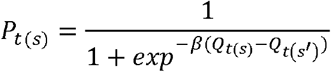

Here, the inverse temperature parameter, *β*, controls the degree to which the probability of the participant choosing shape *s, P*_*t*(*s*)_, is determined by the difference in *Q* values.

This model is over parameterized, the three parameters *ρ, ϕ* and *β* produce very similar effects on asymptotic choice (Figure 1c) and therefore cannot be jointly estimated from participant behaviour. In order to account for changes in behaviour, two of the three parameters have to be fixed while the other (as well as the learning rate) remains free. Doing this is equivalent to making a statement about the presumed cause of the change in behaviour. However, as illustrated in Figure 1 and Table 1, while the three different parameters have the same effect on choice, they act on distinct components of the learning and decision-making process. As a result, it is possible to discriminate between their effects, but doing so requires access to internal model variables such as the expectation and prediction error.

**Table 1:**
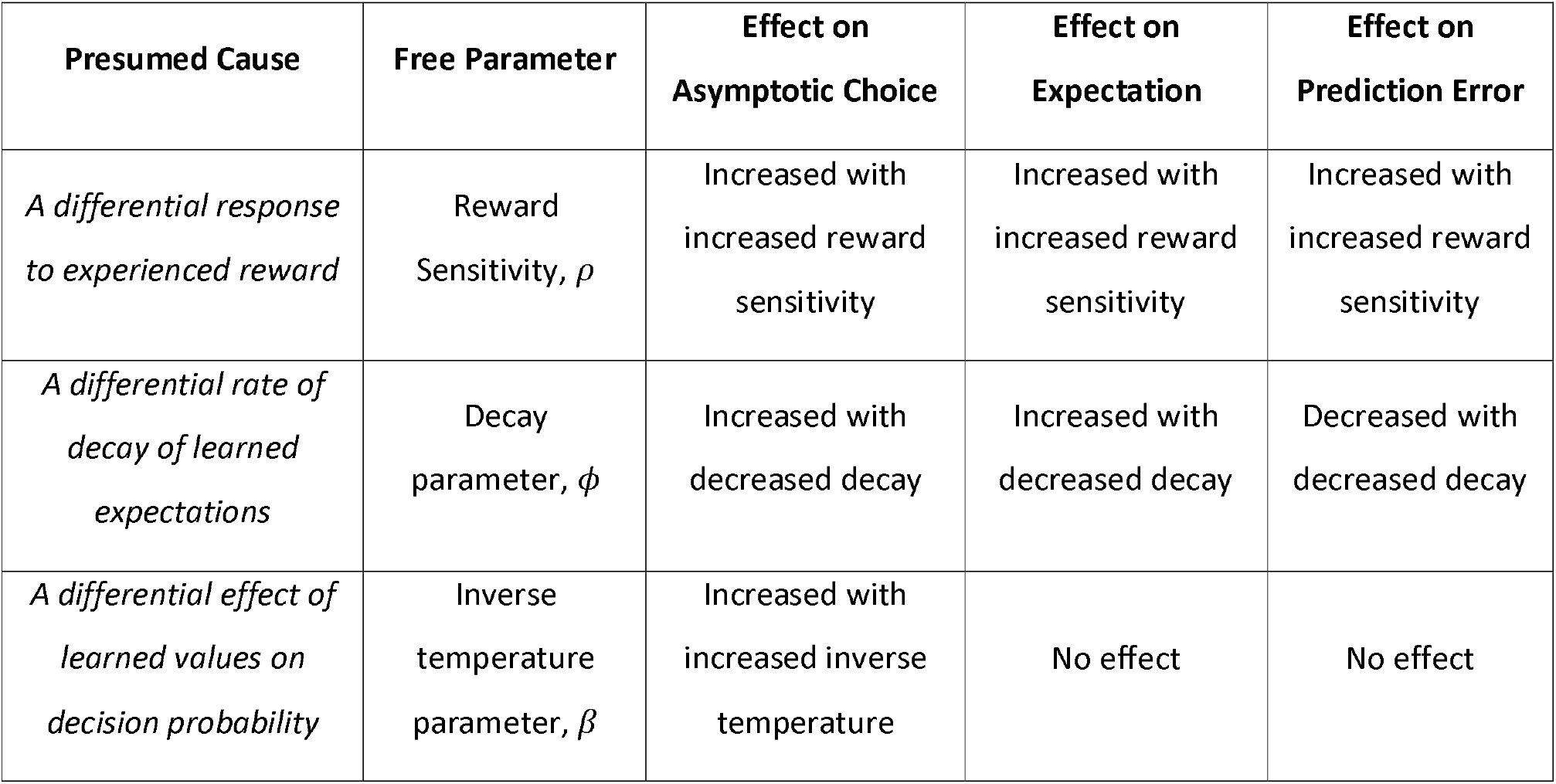
The three causal proposals of changes in asymptotic choice associated with the reward sensitivity, decay and inverse temperature parameters of the reinforcement learning model.

| Presumed Cause | Free Parameter | Effect on Asymptotic Choice | Effect on Expectation | Effect on Prediction Error |
| --- | --- | --- | --- | --- |
| *A differential response to experienced reward* | Reward Sensitivity, $\rho$ | Increased with increased reward sensitivity | Increased with increased reward sensitivity | Increased with increased reward sensitivity |
| *A differential rate of decay of learned expectations* | Decay parameter, $\phi$ | Increased with decreased decay | Increased with decreased decay | Decreased with decreased decay |
| *A differential effect of learned values on decision probability* | Inverse temperature parameter, $\beta$ | Increased with increased inverse temperature | No effect | No effect |

### Model Fitting

In order to fit the three model variants described in Table 1 to participant choice, the joint posterior probability of the free parameters for each variant was calculated for each participant separately, given their choices. Each participant’s parameter values were estimated as the expected value of the marginalised parameter distribution (19,47). *ρ* and *β* parameters were sampled in log space while *α* and *ϕ* parameters were sampled in logit space. All statistical analyses were performed on transformed parameters.

### MRI Image Acquisition

MRI images were acquired using a 3T Siemens Prisma scanner with a 32-channel head-coil. T1-weighted structural images had a voxel resolution of 1 × 1 × 1 mm^3^, echo time of 3.97 msec, repetition time of 1900msec and flip angel of 8°. BOLD Images were T2-weighted echo-planar images. Sixty slices were acquired with a voxel resolution of 2.4×2.4×2.4mm^3^, repetition time of 800msec, echo time of 30msec, flip angle of 52°, field of view of 216mm and the multiband acceleration factor was 6 interleaved. Fieldmaps were collected with echo times of 4.92 and 7.38ms, repetition time of 590msec and flip angle of 46°. Cardiac and Respiratory data were collected and used as denoising regressors.

### MRI Data analysis

MRI data were analysed using FSL (FMRIB Software Library v6.0) tools. Pre-processing involved removal of non-brain structures(48), motion correction(49), spatial smoothing, un-warping using fieldmaps and high-pass temporal filtering (cut-off 60 seconds). Functional images were registered non-linearly to corresponding structural images via a high contrast functional image and BBR (49,50).

Task events were represented using separate explanatory variables for the presentation of stimuli in win and loss trials (2s period during which stimuli were first presented), and separate variables representing the four possible outcomes (2s period during presentation of win, no-win, loss or no-loss outcomes). Additional EVs were included to account for respiratory and cardiac noise. Activity associated with expectation during learning was captured as the relative difference between signal during the stimuli presentation period for win vs. loss condition trials. Post-hoc analyses then compared expectation associated activity between the 1^st^ vs 2^nd^ half of trials in a block (i.e. when expectation should be low relative to when it should be high, Figure 1d). The contrast between ‘win’ and ‘no-win’ outcomes, and ‘no-loss’ and ‘loss outcomes’ were used as simple non-model-based measures of prediction errors. Note that, while this analysis makes no assumption about how participants are learning during the task, it does assume that the observed activity during outcome periods reflects participants’ prediction error rather than just experienced outcome (i.e. the response to an outcome is reduced if that outcome is expected). We therefore supplemented this analysis with a model-based analysis in which the estimated prediction errors from the belief decay model were used as parameter regressors in place of the binary outcome regressors. Model-based results utilizing prediction errors generated using the inverse temperature and reward sensitivity models are reported in the supplementary material.

First-level analyses were run for each participant and both blocks of the task. The outputs of these analyses were then averaged, within subject, across the blocks and entered into a higher-level random-effects analysis which assessed the difference between the two groups. The higher-level analysis was restricted to anatomical ROIs of reward sensitive regions; mPFC, OFC (defined using the Harvard-Oxford Structural Atlas library (51)) and the Ventral Striatum (defined using the Oxford-GSK-Imanova Structural–anatomical Striatal Atlas (52)). Group level inference was performed using the FSL nonparametric permutation tool (Randomise) with 5000 permutations, threshold free cluster enhancement method and family-wise error correction (p<0.05).

## Results

Participants were young, highly educated and evenly split between females and males (Table 1).

### Pramipexole Specifically Increases Asymptotic Choice of Rewarded Stimuli

There was a significant group*valence*time interaction for choice accuracy across behavioural sessions [Figure 2; F(1,38)=10.517 p=0.002]. Win trial accuracy increased after treatment in the Pramipexole group [t(20)=2.347 p=0.029], with no significant change in loss trial accuracy across sessions [t(20)=1.158 p=0.26] and no change in either reward (p=0.86) or loss trial accuracy (p=0.172) in the placebo group. The pramipexole and placebo groups did not differ significantly on reward or loss trial accuracy at baseline (p’s=0.435 and 0.395 respectively) or post-intervention (p’s=0.179 and 0.375 respectively).

**Fig 2.**
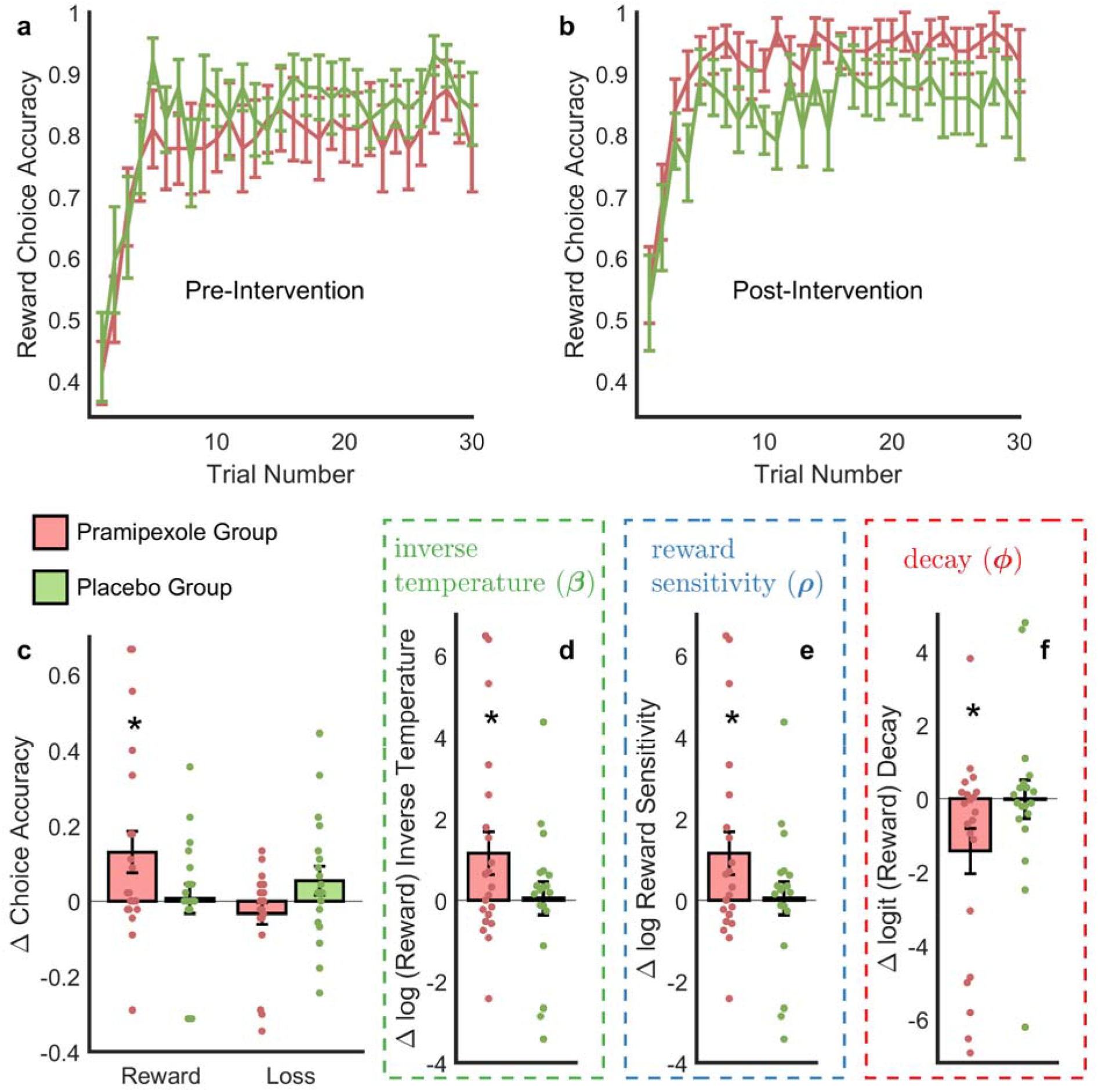
(a-b). Learning curves depicting reward choice accuracy in the (a) pre-intervention and (b) post-intervention session. Green (pink) curves represent the placebo (pramipexole) group. Error-bars represent SEM. The curves represent the proportion of runs in which a participant chose the advantageous shape (i.e. the shape associated with a 70% probability of receiving a ‘win’ outcome) in a given trial. See Figure S3 for loss learning curves. **(c)** Mean (SEM) pre-vs-post intervention change in reward/loss condition choice accuracy. **(d-f)** Three variants of the reinforcement learning model were fitted to participant choice data, in each variant two parameters were fixed and the third (and the learning rate) were allowed to vary. The results show the pre-post intervention change when the reward sensitivity **(d)**, inverse temperature **(e)**, and decay parameter **(f)** were allowed to vary. As can be seen anyone of these three parameters can capture the effect of pramipexole (see Figure S3 for illustration of the degree to which the models are able recapitulate participant learning curves). Green (pink) bars represent the placebo (pramipexole) group. Error-bars represent SEM. Scatter plots overlaying bar graphs depict corresponding individual values. Asterisks represent significant (ps<0.05) pre-vs-post intervention change.

### The Behavioural Effects of Pramipexole May be Attributed to Increased Reward Sensitivity or Choice Stochasticity or to Reduced Reward Value Decay

We fitted participants’ behaviour to the versions of the reinforcement learning model described in the methods section. All three models were able to account for participant behaviour (see Figures S1-3 for model comparison and diagnostics). Specifically, the observed effect of pramipexole may result from increased reward sensitivity (Figure 1d/2e; group * valence * time F(1,38)=5.81 p=0.021), decreased reward value decay (Figure 1e/2f; group * valence * time F(1,38)=7.96 p=0.008) or increased inverse temperature (Figure 1f/2d; group * valence * time F(1,38)=5.81 p=0.021).

### Pramipexole Increases BOLD Signal during Anticipation of Rewards vs. Losses

Anticipation of reward stimuli, as measured using the activity during presentation of stimuli in reward relative to loss trials, was increased in participants receiving pramipexole relative to placebo in the OFC ROI (Peak voxel x=34, y=77, z=31; voxel size:8; p=0.0376) (Figure S4). There were no significant clusters for this contrast in the mPFC or VS ROIs. We next examined the development of win and loss expectations during the task blocks. As illustrated in Figure 1d, expectations develop as learning proceeds. We captured this process by subtracting the response to stimuli in the first half of trials (trial 1-15) from the latter half (trials 16-30) separately for reward and loss trials. Within the reward condition, participants receiving pramipexole had greater increase in activity across the block than those receiving placebo in the OFC ROI (Peak voxel x=30, y=76, z=36; voxel size:57; p=0.019) (Figure 3a-b). There were no clusters for this contrast within mPFC or VS ROIs, nor for the OFC/mPFC/VS ROIs during loss stimulus presentation. These results are consistent with pramipexole causing an increase of reward sensitivity or reduction in reward expectation decay as both processes lead to increased reward expectation (Figure 1d). They are not consistent with an increase in inverse temperature, which does not require a change in expectations.

**Fig 3.**
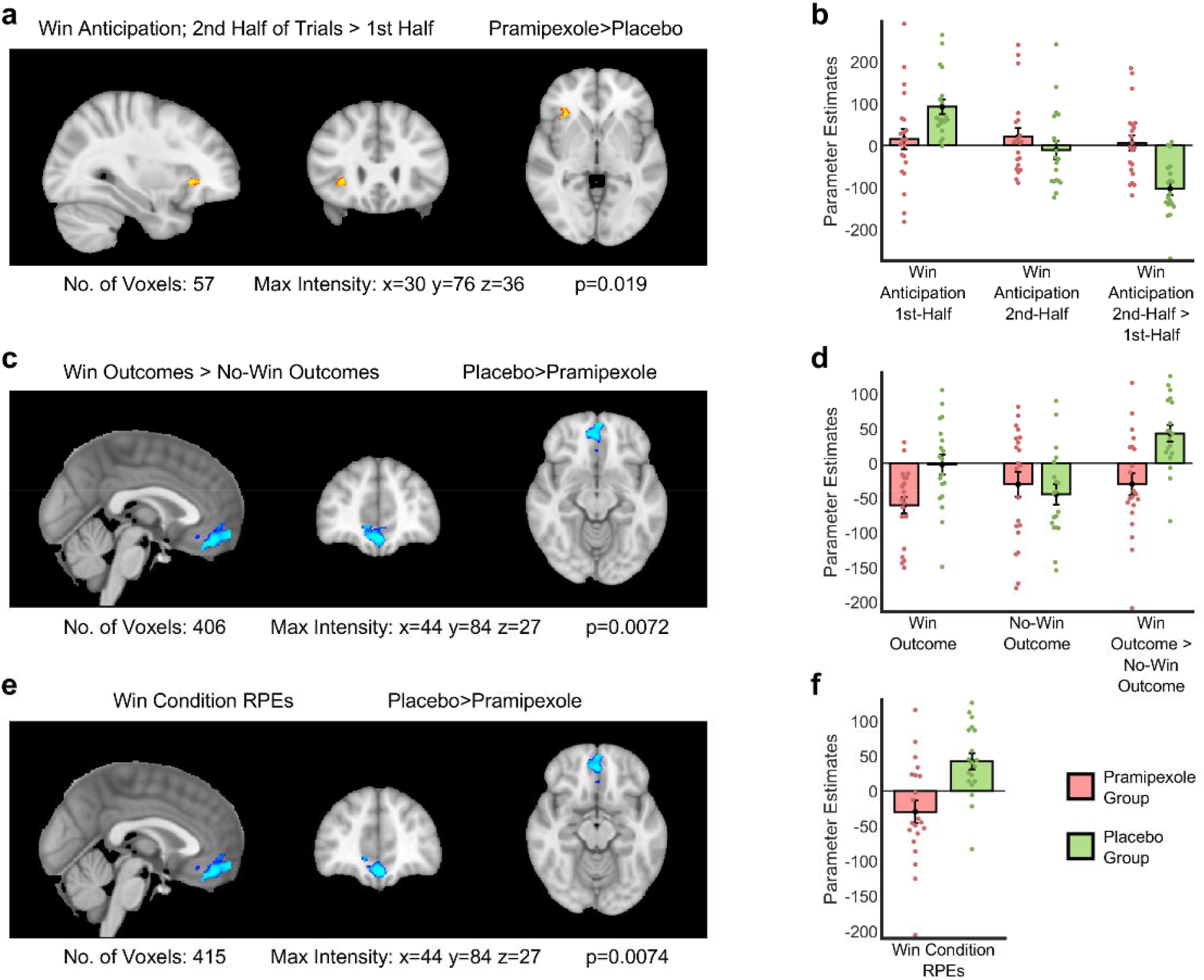
Results of fMRI analyses. **(a)** The (red-yellow) coloured area represents the cluster of significantly increased activity compared to the placebo group in the OFC ROI during win-anticipation in the 2^nd^ half > 1^st^ half of win-condition trials (Peak voxel x=30, y=76, z=36; voxel size:57; p=0.019) and **(b)** parameter estimates extracted from the area of significantly increased activity in 3a associated with win-anticipation in the 1^st^ half, 2^nd^ half, and 2^nd^ half > 1^st^ half of win-condition trials. **(c)** The (blue) coloured area represents the cluster of significantly decreased activity compared to the placebo group in the mFC ROI associated with win-outcomes > no-win-outcomes (Peak voxel x=44, y=84, z=27; voxel size:406; p=0.0072) and **(d)** parameter estimates extracted from the area of significantly increased activity in 3c associated with win- outcomes, no-win-outcomes, and win-outcomes > no-win-outcomes. **(e)** The (blue) coloured area represents the cluster of significantly decreased activity compared to the placebo group for win-condition RPEs in the mFC ROI (Peak voxel x=44, y=84, z=27; voxel size:415; p=0.0074) and **(f)** parameter estimates extracted from the area of significantly increased activity in 3e associated with win-condition RPEs. For 3 a, c and e, areas of significantly increased/decreased activity are threshold free cluster enhancement corrected with a family-wise error cluster significance level of p ≤ 0.05. For 3 b, d and f, green (pink) bars represent the placebo (pramipexole) group. Error-bars represent SEM. Scatter plots overlaying bar graphs depict corresponding individual parameter estimates.

### Pramipexole Decreases the BOLD Signal Associated with Rewarded Prediction Errors

The response to rewarded outcomes, as measured using activity associated with win relative to no-win outcomes, was reduced in the pramipexole group relative to the placebo group in the mPFC ROI (Peak voxel x=44, y=84, z=27; voxel size:406; p=0.0072) (Figure 3c-d). Pramipexole did not influence activity in loss relative to no-loss trials, or in the OFC or VS ROIs. This same effect was apparent using a regressor coding reward prediction errors derived from the decay model (Figure 3e-f; see supplementary materials for summary of analyses using other model variants).

### Questionnaire scores

No effect of drug treatment was found for any of the questionnaire measures, other than the anticipatory subscale of the TEPS, which was driven by a higher baseline score in the pramipexole group (Table 2). Behavioural and neuroimaging analyses controlling for baseline TEPS scores are reported in the supplementary materials.

**Table 2:** Demographic, physical, and psychological characteristics of the Pramipexole and Placebo groups, presented as ‘means (standard deviations)’

|  | **Pramipexole**<br>**(n = 21; 10 male)** | **Placebo**<br>**(n = 19; 10 male)** |
| --- | --- | --- |
| **Age** | 22.5 (3.7) | 24.5 (6.9) |
| **Body mass index** | 22.4 (2.6) | 24.0 (2.9) |
| **Years in full-time education** | 16.8 (2.9) | 17.5 (3.1) |
| **IQ estimate (Spot-the-word Test)** | 108.3 (8.1) | 111.9 (7.6) |
| **Neuroticism (Eysenck Personality Questionnaire)** | 4.2 (3.7) | 4.3 (3.7) |
| **Psychoticism (Eysenck Personality Questionnaire)** | 2.5 (2.1) | 2.8 (2.1) |
| **Extraversion (Eysenck Personality Questionnaire)** | 14.7 (4.5) | 14.5 (3.7) |
| **Lie (Eysenck Personality Questionnaire)** | 9.5 (4.6) | 7.5 (3.4) |
| **Trait Anxiety (State-Trait Anxiety Inventory)** | 31.2 (9.1) | 32.1 (9.1) |
| **Depression at inclusion (Beck Depression Inventory)** | 1.6 (1.7) | 2.5 (4.0) |

**Table 3:** Questionnaire scores before and after the intervention: Becks Depression Inventory (BDI), Befindlichkeitsskala (BFS), Positive and Negative Affect Schedule (PANAS), State-Trait Anxiety Inventory (STAI), Snaith-Hamilton Pleasure Scale (SHAPS), Temporal Experience of Pleasure Scale (TEPS), Oxford Happiness Questionnaire (OXH) and Impulsive-Compulsive Disorders in Parkinson’s Disease-Rating Scale (QUIP). Scores are presented as ‘means (standard deviations)’. Asterisks represent significant between-group, within-condition difference in questionnaire scores.

|  | Pramipexole |  | Placebo |  | Group*Time Interaction |
| --- | --- | --- | --- | --- | --- |
|  | Baseline | On drug | Baseline | On drug |  |
| **BFS total** | 11.7 (12.4) | 15.4 (14.7) | 13.8 (11.2) | 14.8 (14.2) | p=0.44 |
| **BFS energy** | 4.0 (5.0) | 4.9 (5.8) | 3.5 (3.5) | 4.7 (5.7) | p=0.87 |
| **BFS mood** | 7.8 (8.8) | 10.5 (10.1) | 10.4 (8.3) | 10.1 (10.3) | p=0.17 |
| **STAI state** | 28.9 (6.5) | 27.8 (5.4) | 28.6 (6.1) | 29.1 (5.2) | p=0.34 |
| **BDI** | 1.9 (2.5) | 2.9 (3.4) | 2.8 (3.3) | 2.7 (3.3) | p=0.26 |
| **PANAS positive present** | 36.4 (7.7) | 34.5 (8.3) | 32.5 (8.6) | 32.6 (7.9) | p=0.25 |
| **PANAS negative present** | 11.2 (1.5) | 11.0 (1.5) | 11.3 (1.3) | 11.8 (2.7) | p=0.15 |
| **PANAS positive today** | 36.8 (8.0) | 34.8 (9.1) | 32.9 (8.2) | 32.3 (8.1) | p=0.46 |
| **PANAS negative today** | 11.4 (2.0) | 11.4 (1.7) | 11.6 (1.7) | 11.8 (2.4) | p=0.68 |
| **PANAS positive last week** | 37.7 (9.2) | 37.0 (7.7) | 34.2 (7.6) | 34.6 (9.3) | p=0.61 |
| **PANAS negative last week** | 13.3 (2.8) | 12.7 (3.2) | 14.1 (4.1) | 12.6 (2.7) | p=0.44 |
| **SHAPS** | 0.5 (1.0) | 0.7 (1.6) | 0.2 (0.7) | 0.3 (0.7) | p=0.70 |
| **TEPS total** | 85.3 (10.0) | 82.1 (9.5) | 79.3 (8.2) | 79.2 (10.0) | p=0.16 |
| **TEPS anticipatory** | 47.1 (6.1)* | 44.6 (5.5) | 42.1 (4.9)* | 42.3 (5.4) | **p=0.04** |
| **TEPS consummatory** | 38.1 (5.6) | 37.5 (5.9) | 37.3 (4.4) | 36.8 (5.4) | p=0.83 |
| **OXH** | 134.8 (19.2) | 138 (18.4) | 132.4 (19.6) | 132 (19.7) | p=0.10 |
| **QUIP** | 12.3 (8.3) | 8.7 (8.5) | 16.6 (11.2) | 13.8 (10.4) | p=0.72 |

## Discussion

Sustained treatment with pramipexole increases the asymptotic choice of rewarded stimuli while simultaneously enhancing neural activity during the anticipation of rewarded trials and suppressing the response to win outcomes. This indicates that it enhances reward learning by reducing the decay of value estimates and suggests a cognitive mechanism of action by which it may ameliorate the reduced reward learning characteristic of depression and anhedonia.

Pramipexole specifically increased asymptotic choice of highly rewarded stimuli, with no effect on the choice of stimuli associated with loss. This is in contrast with the majority of previous experimental studies of pramipexole which have found that it impairs reward learning (30–33). However, these studies have generally administered a single, low dose of pramipexole which is thought to act primarily on inhibitory, dopamine presynaptic autoreceptors (40,41). The two-week administration used in the current study was selected to avoid this effect and to assess the more clinically relevant action of the drug on post-synaptic receptors. The current results are consistent with the few studies of patient groups in which longer treatment durations were used, and which also found an increase in reward choice following pramipexole treatment (36,53). Together, these studies indicate that putatively post-synaptically acting, sustained dosing of pramipexole acts to enhance reward learning. Depression is associated with reduced reward learning (5–16). Overall, therefore, the impact of pramipexole on learning is opposite to that associated with depression and is consistent with the antidepressant effects of the drug (27–29). The purported underlying mechanism (reduced reward expectation decay) is compatible with reports, by patients suffering from pramipexole-induced compulsive disorders, of persistent pre-occupation with rewarding activities even in the absence of obvious cues (54).

The increase in asymptotic reward choice following treatment with pramipexole may be produced by a number of distinct cognitive mechanisms (Figure 1). It is not possible to arbitrate between these using choice behaviour from the PILT task alone, rather estimates of internal model processes are required. We used functional neuroimaging of reward-sensitive neural areas during the presentation of task stimuli and the receipt of outcomes to provide estimates of these internal processes. Pramipexole was found to increase anticipatory activity during rewarded trials in the OFC, a region in which activity commonly tracks expected value (55–57) and in which activity is found to be altered in depression (58,59). Pramipexole also reduced the response to win outcomes and reward prediction errors in the mPFC, a node in a previously described, positive-valence-specific, reward prediction error network (60). Contrary to our hypothesis, this pattern of effect suggests that pramipexole enhances value expectations, and therefore reward learning, by reducing the decay of value estimates between trials rather than by enhancing the effective value of the outcomes. Depression itself is associated with a reduced BOLD response to rewarding outcomes (7,21–24), which suggests that the reduced learning in patients (5–16) is the result of a lower effective value of rewards rather than a difference in decay of value estimates. The current findings indicate that pramipexole does not act directly to reverse the cognitive profile of depressed patients, but rather improves reward learning via a separate mechanism. This result may go some way to explaining why the clinical response to pramipexole in depression seems to be higher in patients with intact, rather than impaired, baseline reward learning (34). Specifically, as pramipexole does not increase reward sensitivity, the impact of the drug on reward learning, and presumably on symptoms of depression, will depend on an intact response to rewarding outcomes, and will be reduced in those patients with an impaired response. In other words, there is little point in decreasing the decay of reward value estimates if these estimates have been systematically lowered by reduced reward sensitivity. This interpretation raises the question of whether alternative approaches to enhancing reward learning, such as kappa-opioid receptor antagonism (61) or cognitive interventions (62), might act to enhance reward sensitivity and whether the effects of these treatments may therefore be complementary to those of pramipexole.

An outstanding question raised by the current results is how pramipexole might act to reduce the decay of estimated values. One possibility is that this effect is related to the role of dopamine in working memory (63). Previous modelling work has demonstrated that simple learning tasks are often solved using a mixture of working memory and reinforcement learning based processes, with working memory acting to reduce prediction error responses by maintaining distinct representations of current value (20). The observed effect of pramipexole in this study may therefore reflect an increase in the degree to which participants rely on working memory when completing the PILT. However, a general enhancement of working memory should also influence loss learning, rather than produce a reward-specific effect as found here. It is therefore necessary to evoke some form of valence-specific working memory effect to explain the current findings. Ultimately, the potential role of working memory in the effect of pramipexole would best be tested by manipulating memory load during learning (17). An alternative explanation for the reduced decay in estimated values, that may more naturally incorporate valence specificity, is that value estimates may be rationally combined with prior beliefs during learning, and pramipexole may change these underlying implicit beliefs. By this view, individuals maintain a global estimate of the likelihood of experiencing positive events and they use this estimate to moderate their local estimate of reward value during the task. If, for example, an individual’s global estimate is pessimistic, with positive events judged to only occur rarely, it will act to particularly reduce the estimated value of highly rewarded stimuli. The effect of pramipexole may therefore be understood as inducing a more positive global estimate of the likelihood of rewarding events, which reduces the degree to which local estimates are downgraded. If correct, this would suggest that pramipexole should also act to increase other measures of optimism bias (64).

The current study has a number of limitations. Most obviously, the population recruited were non-clinical healthy participants. A non-clinical population was selected to reduce phenotypic variation among participants and thus enhance the sensitivity of this experimental medicine study to detect the pharmacological effects of pramipexole. However, this design is not able to assess the degree to which change in reward learning mediates clinical response in patients. Answering this question requires a clinical trial of pramipexole in which patients complete the PILT task before and after initiating treatment with pramipexole or placebo. We are currently undertaking such a trial (65).

A 2-week course of pramipexole enhanced asymptotic choice of highly rewarded stimuli, while reducing the neural response to rewarding outcomes. These results indicate that pramipexole enhances reward learning by reducing the decay of learned value estimates and suggests a potential cognitive mechanism by which it may act to ameliorate symptoms of depression.

## Supporting information

Supplementary Methods and Results

## Data Availability

All data produced in the present study are available upon reasonable request to the authors

## Acknowledgements

This study was supported by the Oxford Health NIHR Biomedical Research Centre and the Medical Research Council. The Wellcome Centre for Integrative Neuroimaging is supported by core funding from the Wellcome Trust (203139/Z/16/Z). MB, MM and CJH are supported by the Oxford Health NIHR Biomedical Research Centre. MB is supported by the NIHR Oxford Cognitive Health Clinical Research Facility. The views expressed are those of the authors and not necessarily those of the Wellcome Trust, the Medical Research Council, the NHS, the NIHR, or the Department of Health. None of these bodies had a significant role in the design, collection and analysis of data, or the decision to publish this article.

