## Supplementary Methods and Results for "Pramipexole Enhances Reward Learning by Preserving Value Estimates"

**Model Selection**

We utilizing reinforcement learning models as outlined in table 1 and the body of the methods section of the paper. In brief, the three model-types respectively utilized reward sensitivity, decay and inverse temperature parameters to alter asymptotic choice behaviour. All models utilized learning rate parameters to update the valuation of task stimuli. For each model type we tested whether participants utilized different parameter values for each of the two conditions or whether they used the same parameter values across both conditions. We also tested whether participants updated stimulus-values reciprocally, i.e. whether they updated the unchosen stimulus by the counterfactual outcome in a given trial. We expected that they would do so as we instructed participants that stimulus-outcome contingencies for individual trials would be reciprocal. We initialized stimulus values at 0.5 i.e. the midway point between the values of possible outcomes (0 and 1). We tested 4 variants of each model-type, totaling 12 models tested as outlined in table S1.

| **Model-Type** | **Model** | **Parameters** | **Value-Updating** |
| --- | --- | --- | --- |
| Decision Determinacy Models | 1 | 2 parameters: α, β | Non-Reciprocal Updating |
|  | 2 | 4 parameters: α_win_, α_loss_, β_win_, β_loss_ | Non-Reciprocal Updating |
|  | 3 | 2 parameters: α, β | Reciprocal Updating |
|  | 4 | 4 parameters: α_win_, α_loss_, β_win_, β_loss_ | Reciprocal Updating |
| Reward Sensitivity Models | 5 | 2 parameters: α, ρ | Non-Reciprocal Updating |
|  | 6 | 4 parameters: α_win_, α_loss_, ρ_win_, ρ_loss_ | Non-Reciprocal Updating |
|  | 7 | 2 parameters: α, ρ | Reciprocal Updating |
|  | 8 | 4 parameters: α_win_, α_loss_, ρ_win_, ρ_loss_ | Reciprocal Updating |
| Belief Decay Models | 9 | 2 parameters: α, φ | Non-Reciprocal Updating |
|  | 10 | 4 parameters: α_win_, α_loss_, φ_win_, φ_loss_ | Non-Reciprocal Updating |
|  | 11 | 2 parameters: α, φ | Reciprocal Updating |
|  | 12 | 4 parameters: α_win_, α_loss_, φ_win_, φ_loss_ | Reciprocal Updating |

**Table S1. Models tested.**

We wanted, ultimately, to test which parameter (decision determinacy, reward sensitivity and decay) best accounted for the changes seen in the pramipexole group from the pre-to-post intervention session. However, we first set out to determine which ‘structure’ of RL model to use (i.e. condition-specific vs independent parameters; reciprocal vs non-reciprocal updating). Our a priori belief was that participants would utilize separate parameter values for the two conditions (as reflected in the hypothesis stated in the paper). We believed so because, in our clinical experience, pramipexole appears to alter reward-related decision making seemingly independently of punishment-related decision making. We also believed a priori that participants update stimulus values reciprocally as we instructed them that in any given trial, one shape would yield a reward/punishment while the other would not.

We first compared model structures by averaging BICs (Figure S1a) and AICs (Figure S1b) for each structure across parameter-categories (reward sensitivity/decay/inverse temperature). Those models that utilized condition-specific parameters performed better than those that didn’t and reciprocally updating models performed better than the non-reciprocally updating models. Our AIC/BIC results confirmed our a priori belief that participants used separate, condition-specific, parameters and that they updated values reciprocally.

We then compared BICs (Figure S1c) and AICs (Figure S1d) for each specific model (reward sensitivity i.e model 8/decay i.e. model 12/inverse temperature i.e. model 4) within the winning structure-category. As can be seen, these latter three AICs/BICs are comparable, though the decay parameter model (12) does have higher AIC/BIC scores (mean AIC=276; mean BIC=281) than the inverse temperature and reward sensitivity models (mean AICs=260; mean BICs=274).

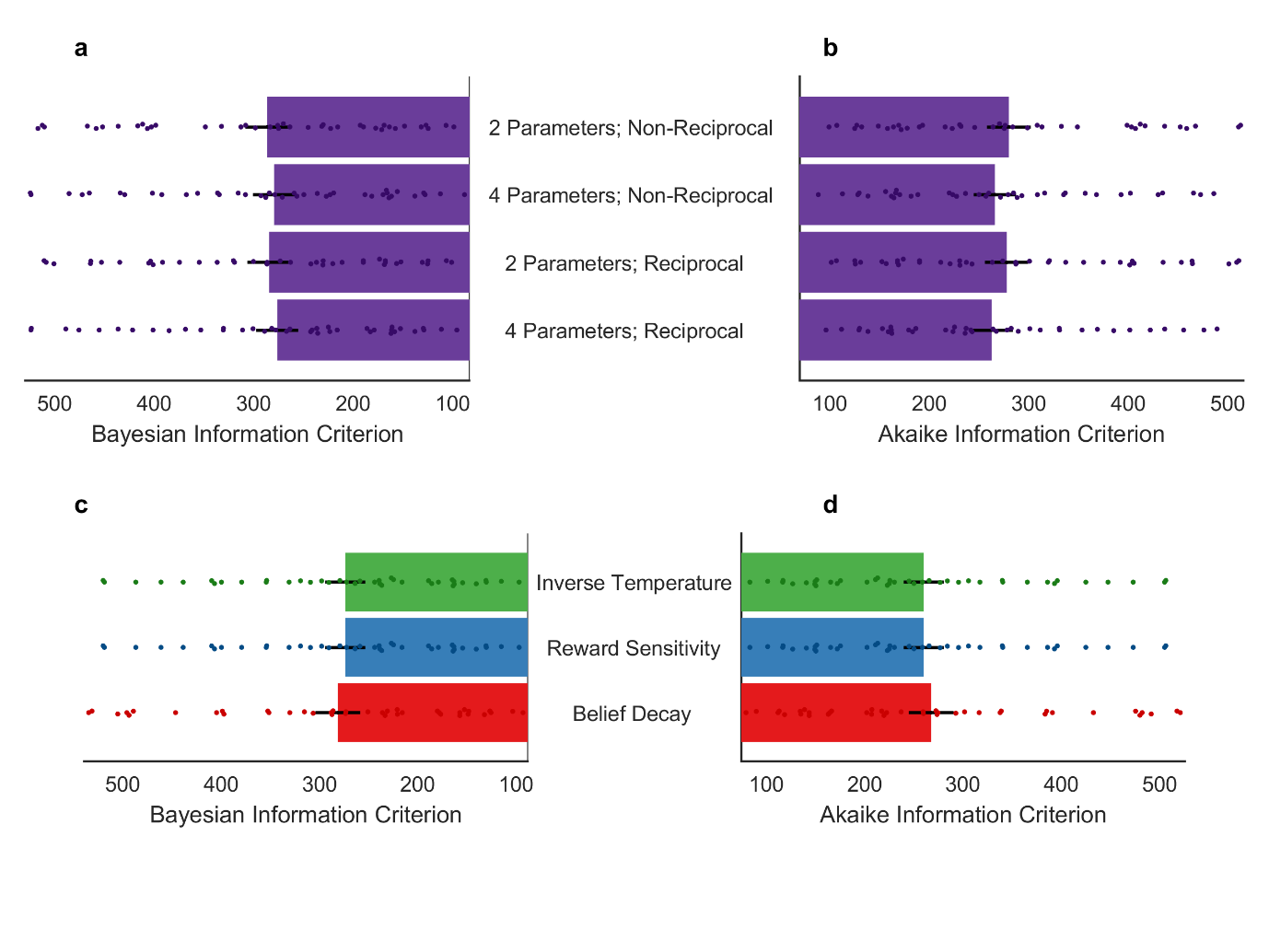

**Figure S1. Average (a) BIC and (b) AIC scores for each model structure:** 2 parameter models with non-reciprocal updating (models 1, 5 and 9), 4 parameter models with non-reciprocal updating (models 2, 6 and 10), 2 parameter models with reciprocal updating (models 3, 7 and 11), 4 parameter models with reciprocal updating (models 4, 8 and 12) (see table S1 for model descriptions)**. Average (c) BIC and (d) AIC scores for each model within the winning structure-category** i.e. the reciprocally updating 4 parameter model that utilised inverse temperatures (model 4) reward sensitivity parameters (model 8) and decay parameters (model 12). Smaller AIC/BIC scores indicate a better model fit. AIC/BIC scores were calculated by the summing across the 3 task blocks and across both sessions. Bars represent mean (SEM) AIC/BIC scores across all participants. Scatter plots overlaying bar graphs depict corresponding individual AIC/BIC values.

As the most pertinent change across sessions was the increase in reward choice accuracy within the pramipexole group, we next assessed how accurately the models replicated this change. We generated reward-condition choices using each model and individual participants’ parameter values for each session, for each respective model. As we did for the model-free analysis in the paper, we used the proportion of ‘accurate’ choices in the latter half of each block. We plotted the change in this metric across sessions for each model (Figure S2). Models that utilized condition-specific parameters appear to perform better than those that utilize condition-independent parameters. There appears to be little difference between those models that update values reciprocally and those that update values non-reciprocally. This analysis therefore supports our a priori belief regarding utilizing condition-specific parameter values but does little to discriminate between reciprocally vs non-reciprocally updating models.

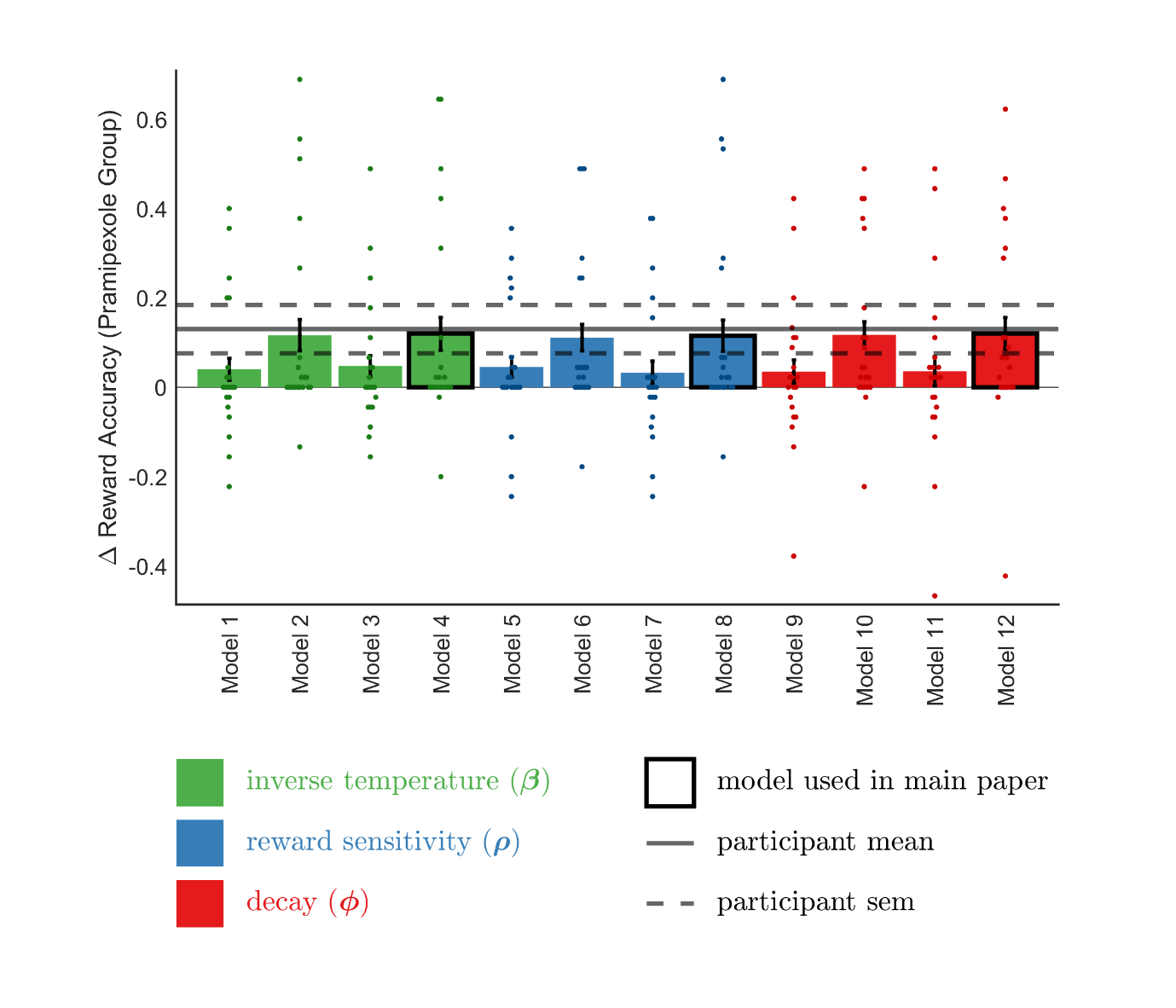
**Figure S2.** **Bar graphs depicting change in model generated choice accuracy using** **pre-vs-post intervention, pramipexole group derived parameters (see table S1 for model descriptions).** Bars represent mean (SEM) change in the reward-accuracy of model-generated choices utilising parameters values inferred for participants that took pramipexole. ‘Decision determinacy’ model results are coloured green, reward sensitivity models blue and decay models red. Models used in the paper are framed in black. Scatter plots overlaying bar graphs depict corresponding individual values.

Taking trial-wise choices generated by each model using individual participants’ parameters, we averaged these model-generated choices within group and condition to create learning curves for each model (equivalent to participants’ learning curves in Figure 2a/b of the main paper). We plotted these learning curves against pramipexole/placebo-group participants’ actual learning curves and visually assessed the fidelity of the model-generated learning curves to participants’ learning curves. We created (2 groups * 2 sessions * 12 models =) 72 plots; it is impractical to present all 72 plots. In brief, the learning curves of those models that utilized condition-specific parameter appeared to have greater fidelity to participant learning curves than those that didn’t. Little separated the apparent fidelity of learning curves generated using models that updated values reciprocally vs non-reciprocally. For illustrative purposes we have presented below 4 plots depicting participant learning curves vs those generated by the 3 models used in the paper (Figure S3). As can be seen, the 3 models used in the paper appear to have comparable fidelity to the participant behaviour though, arguably, the decision determinacy model (model 4) and reward sensitivity model (model 8) generated learning curves appear to have slightly higher fidelity to some of the participant learning curves than does the decay model (model 12) generated learning curves.

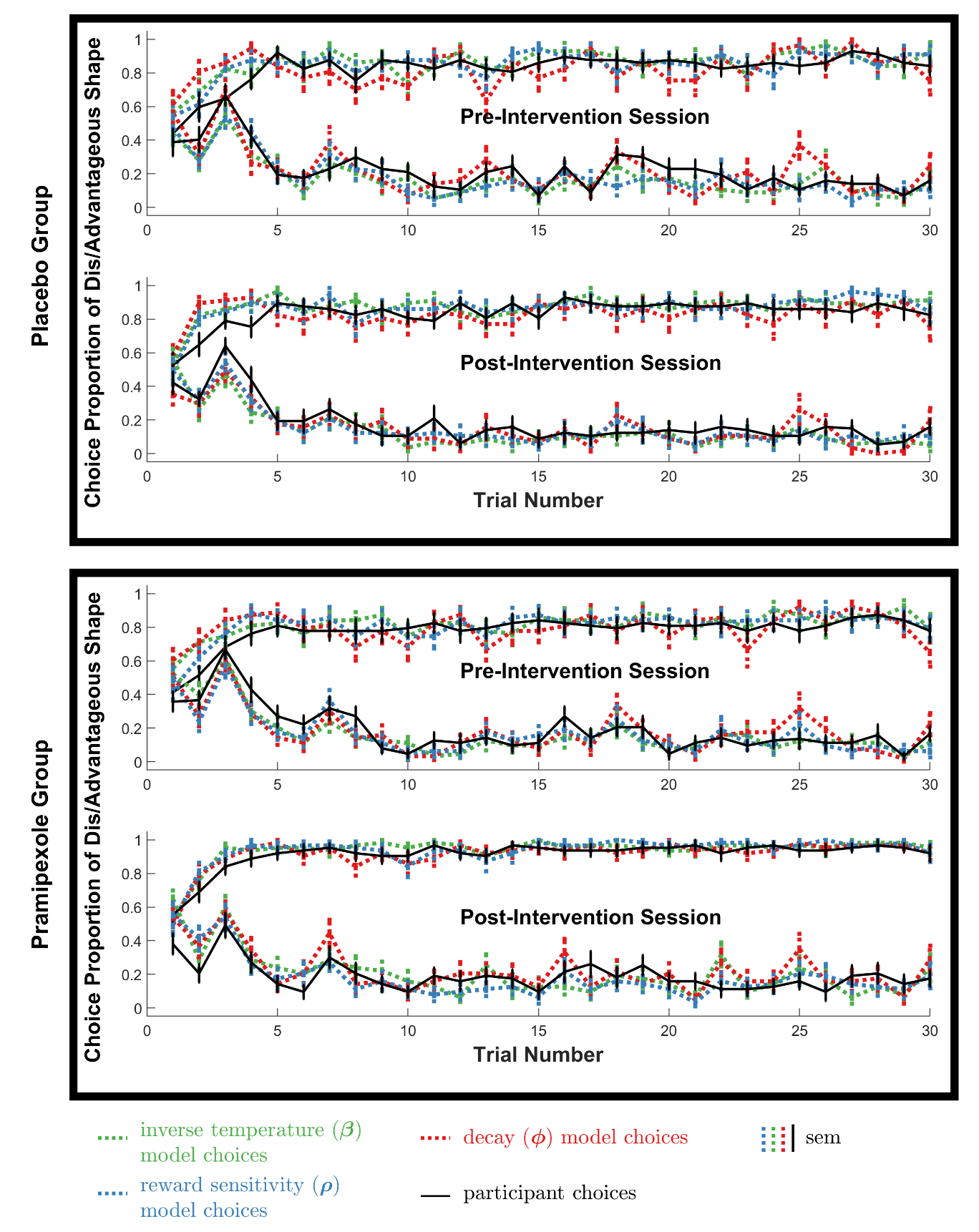

**Figure S3. Learning curves depicting model vs participant choice accuracy in each session.** Approximately concave plots represent reward condition learning curves, that is, time series’ depicting the proportion of participants/simulated-participants that chose the advantageous reward-condition stimulus in a given trial. Approximately convex plots represent loss condition learning curves, that is, time series’ depicting the proportion of participants/simulated-participants that chose the disadvantageous loss-condition stimulus in a given trial. Solid black lines represent participant choices while dotted lines represent model-generated choices. Choices generated by model 4 are coloured green, choices generated by model 8 are blue and choices generated by model 12 are red. Error-bars represent SEM.

In summary, we used positive and negative metrics to assess 12 models comprising 4 versions (reciprocal vs non-reciprocal updating; condition-specific vs independent parameter values) of models that each used one of three different parameter-types (decision determinacy/reward sensitivity/decay) along with learning rate parameters. Models containing condition specific parameters performed better than those that didn’t. Reciprocally updating models performed marginally better than non-reciprocally updating models. We therefore used models 4, 8 and 12 (see Table S1) in the paper. Comparing models 4, 8 and 12, the decision determinacy model (4) and reward sensitivity model (8) had lower mean AIC and BIC scores the decay model (12). All three models re-produce the effect of pramipexole on reward accuracy to a comparable degree. All three models also reproduce participant learning curves to a comparable degree; though, arguably, Models 4 and 8 do so with marginally higher fidelity than model 12 with respect to some of the learning curves.

**Supplementary Results**

**There are no significant reward/loss accuracy differences between the pre-intervention behavioural session and imaging session:** The main behavioural finding presented in the paper was a significant group*valence*(pre vs post intervention behavioural )session interaction for choice accuracy. We likewise compared the pre-intervention behavioural session vs the post-intervention imaging session and found no such group*valence*(pre-intervention vs imaging )session interaction for choice accuracy [F(1,38)=10.517 p=0.096] nor any significant changes in reward/loss condition accuracy across sessions in either group (all ps≥0.086.) We found no significant group*valence*(pre-intervention vs imaging )session interactions for reward sensitivity, decay or decision determinacy parameters (all ps≥0.309), nor any significant changes in win/loss trial parameter values across sessions in either group (all ps≥0.087.)

**The pattern of the main behavioural findings is unchanged by including all (as opposed to the second half of) trials:** In the paper we calculate model-free behavioural results using participants’ choice accuracy in the second half of trials as the focus of the analysis is the effect of pramipexole on the asymptotic choice accuracy. If we instead includr all trials in the analysis, the pattern of results remains the same. The group*valence*session interaction remains significant [F(1,38)=7.572 p=0.009]. The increase in pramipexole group reward condition accuracy across sessions remains significant [t(20)=2.705 p=0.014)]. The change in pramipexole group loss condition accuracy across sessions, as well as the change in placebo group reward and loss accuracy remain non-significant (all ps≥0.172).

**The effect of adding baseline TEPS anticipatory subscale scores as a covariate to the analyses reported in the paper:** By chance, the two groups differed significantly in the anticipatory subscale of the TEPS questionnaire at baseline. We added individuals’ baseline TEPS anticipatory subscale scores to behavioural and significant model free fMRI analyses.

Adding Baseline anticipatory TEPS sub-scores as a covariate, the group*condition*(behavioural )session interaction for remained significant: F(1,37)=8.59 p=0.006. Likewise, the group*condition*(behavioural )session interaction for computational parameter values remains significant: Inverse temperature parameter [F(1,37)=5.521 p=0.024], reward sensitivity parameter [F(1,37)=5.521 p=0.024] and decay parameter [F(1,37)=6.332 p=0.016].

Interestingly, adding individuals’ baseline TEPS anticipatory scores as a co-variate yielded a significant group*condition*session interaction for choice accuracy (F(1,38)=4.457 p=0.042).

The group*condition*(pre-intervention vs fMRI) session interactions for computational parameter values (inverse temperature parameter, reward sensitivity parameter and decay parameters) remain non-significant (ps≥0.167).

Adding baseline TEPS anticipatory sub-scale scores as a co-variate in the analysis of the imaging results reported in the paper, we find that the Win Anticipation > Loss Anticipation; Pramipexole > Placebo and Win Anticipation 2^nd^ Half > 1^st^ Half; Pramipexole > Placebo contrasts are no longer significant. The Win Out > No-Win Out; Placebo > Pramipexole contrast remains significant but the area of significant activity reduces to 5 voxels (Peak voxel x=46, y=81, z=25; voxel size:5; p=0.0434). Likewise the Win Condition RPE; Placebo > Pramipexole contrast remains significant but area of significant activity reduces to 3 voxels (Peak voxel x=46, y=81, z=25; voxel size:3; p=0.0476).

**Model Free fMRI Image: Win Anticipation > Loss Anticipation**

As stated in the paper, activity during win anticipation > loss anticipation was increased in participants receiving pramipexole relative to placebo, in the OFC ROI (Peak voxel x=34, y=77, z=31; voxel size:8; p=0.0376). We present this result below (Figure S4).

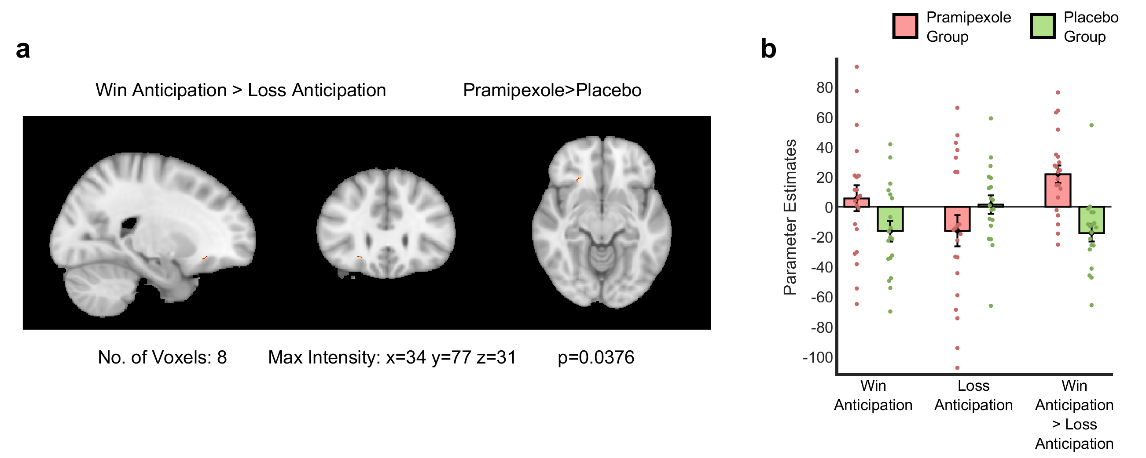

**Fig S4. Model-Free fMRI results (a)** The red-yellow coloured area represents the cluster of significantly increased activity in the pramipexole > placebo group in the OFC ROI during win-anticipation > loss-anticipation (Peak voxel x=34, y=77, z=31; voxel size:8; p=0.0376). Areas of significantly increased activity are threshold free cluster enhancement corrected with a family-wise error cluster significance level of p ≤ 0.05. **(b)** Parameter estimates extracted from the area of significantly increased activity in *S4a* associated with win-anticipation, loss-anticipation and win-anticipation > loss-anticipation.

**Model-Based fMRI results**

As stated in the methods section of the paper, we extracted trial-wise RPEs for each model used in the paper, for each participant, using that participant’s model parameter estimates. We then used those RPEs (demeaned) as regressors in three (decision determinacy, reward sensitivity and decay) model-based analyses. In the paper we present the between-group contrasts for win condition RPEs for the decay model. Below we present between-group contrasts for win-anticipation > loss-anticipation (Figure S5) and win-condition RPEs (Figure S6) from each model-based analysis. Each showed an increased expectation signal by the pramipexole > placebo group in the win > loss condition and showed decreased RPEs by the pramipexole > placebo group in the win condition. All three model-based analyses, therefore, reveal a pattern of BOLD signal that is consistent (only) with the decay model.

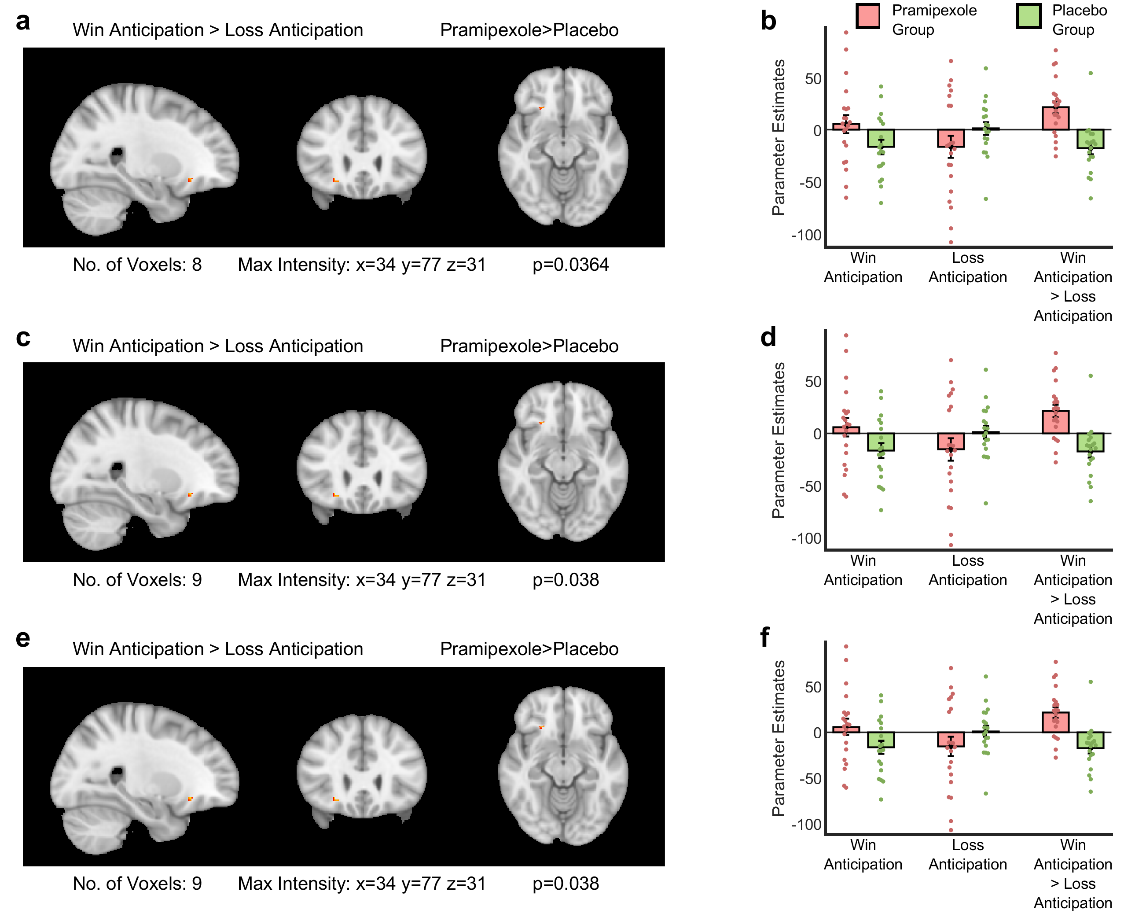

**Fig S5. Model based fMRI (expectation) results: (a,c,e)** Win-anticipation > loss-anticipation contrast from the **(a)** decay-model based analysis **(c)** beta-model based analysis and **(e)** rho model based analysis. The red-yellow coloured areas represent the cluster of significantly increased activity in the pramipexole > placebo group in the OFC ROI. Areas of significantly increased activity are threshold free cluster enhancement corrected with a family-wise error cluster significance level of p ≤ 0.05. **(b,d,f)** Parameter estimates extracted from the area of significantly increased activity in *S5 a, c* and *e* respectively, associated with win-anticipation, loss-anticipation and win-anticipation > loss-anticipation. Green (pink) bars represent the placebo (pramipexole) group. Error-bars represent SEM. Scatter plots overlaying bar graphs depict corresponding individual parameter estimates.

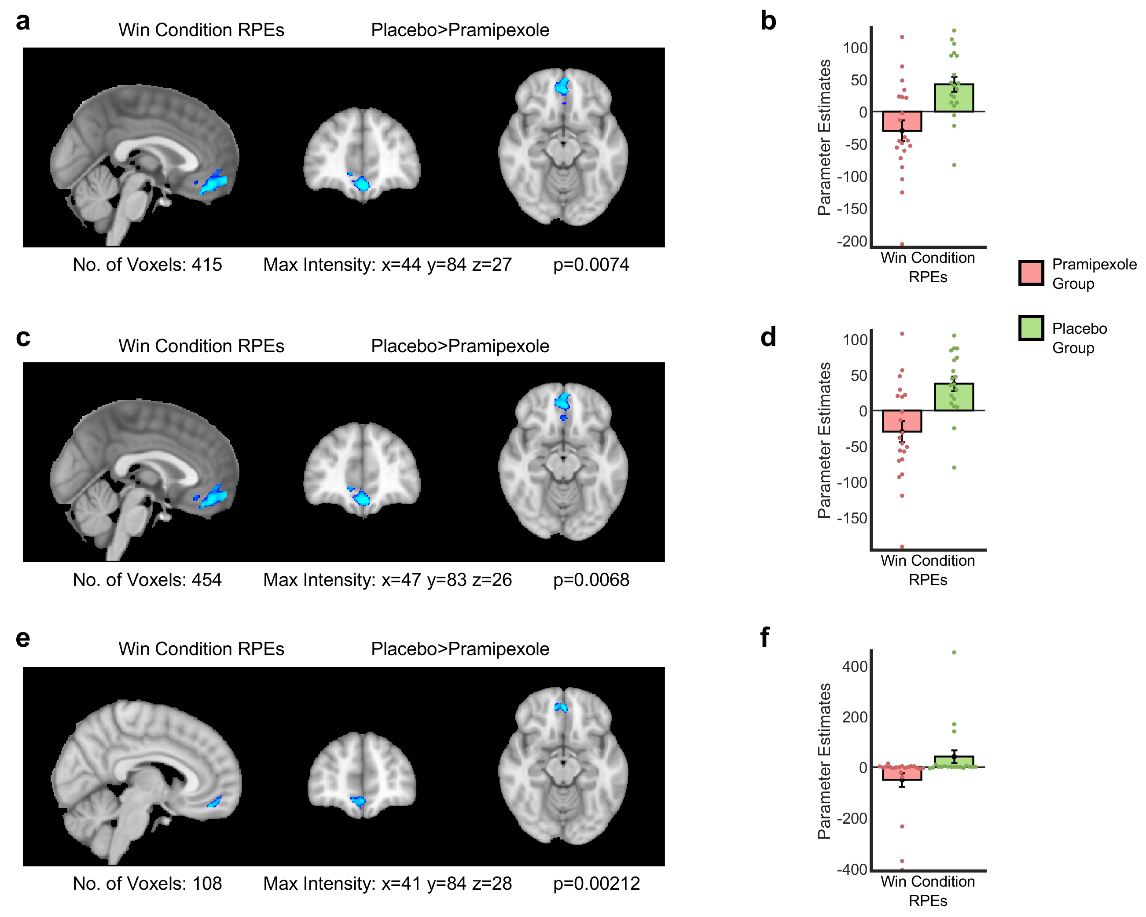

**Fig S6. Model based fMRI (expectation) results: (a,c,e)** Win-condition RPE contrasts from the **(a)** decay-model based analysis **(c)** beta-model based analysis and **(e)** rho-model based analysis. The (blue) coloured area represents the cluster of significantly decreased in the pramipexole > placebo group for win-condition RPEs in the mFC ROI. Areas of significantly decreased activity are threshold free cluster enhancement corrected with a family-wise error cluster significance level of p ≤ 0.05. **(b,d,f)** Parameter estimates extracted from the areas of significantly increased activity in *S6 a, c* and *e* respectively, associated with win-condition RPEs. Green (pink) bars represent the placebo (pramipexole) group. Error-bars represent SEM. Scatter plots overlaying bar graphs depict corresponding individual parameter estimates.
